# A Practical Introduction to Wavelet Analysis in Electroretinography

**DOI:** 10.1101/2025.07.25.25331915

**Authors:** Yousif Shwetar, David Lalush, Jason McAnany, Brett Jeffrey, Melissa Haendel

## Abstract

**Purpose:** To provide a conceptual understanding of the continuous and discrete wavelet transforms (CWT, DWT) for clinical electroretinography (ERG) analysis, and how these methods uncover time-frequency features that complement traditional time-domain analysis.

**Methods:** A technical overview without the use of mathematical formula describing the basics of CWT and DWT and implementation considerations. We also review an example of four standard ISCEV ERG recordings from a healthy male (between 30-34 years of age) and a male (between 15-19 years of age) with complete congenital stationary night blindness (CSNB).

**Results:** Wavelet analysis uncovered time-frequency signatures absent in raw traces. In light-adapted flicker, the normal ERG showed a ∼30 Hz response with harmonics up to 90 Hz, whereas CSNB was largely attenuated. For LA 3 and dark-adapted flashes, normal CWTs concentrated energy < 100 Hz between 0.04-0.08 s, while CSNB demonstrated lowered or almost absent energy profiles in comparison. DWT indices exhibited a similar pattern, with normal recordings demonstrating high energy responses early in the 7, 15, and 29 Hz frequency bands, while CSNB registered markedly lower values.

**Conclusions:** CWT and DWT provide complementary and objective insight into ERG responses. Open-source MATLAB toolkit and step-by-step tutorial provided herein lower technical barriers and enable use by the broader community.

## Introduction

The electroretinogram (ERG) measures the change in voltage across the retina in response to a light stimulus, with clinical ERG analysis typically performed in the time domain (e.g., amplitude, implicit time, waveform shape) [1, 2]. However, the ERG is a complex signal with contributions from multiple cellular sources, and conducting analysis purely in the time-domain may limit additional valuable insight that time-frequency based metrics can offer. Several studies have demonstrated the value of time-frequency domain analysis, particularly through wavelet transforms (WTs) [3–19]. For example, Gauvin and colleagues applied wavelet analysis to isolate the ON and OFF components of the photopic ERG across a range of recording protocols (variations in stimuli strength and duration), demonstrating a clear relationship between associated wavelet-derived descriptors and the functional contributions of ON- and OFF-bipolar cells [3]. In another study of theirs, they used a similar approach to measure the relative energy of the oscillatory potentials (OPs) in the photopic ERG, and found that certain pathological conditions exhibit characteristic energy contributions in these high-frequency oscillations [4]. WTs are objective, not based on a single time point in the waveform, can be resistant to the effects of noise, and have quick computational implementation. However, the technique has not been widely adopted in the field to date, in part due to perceived challenges in implementation.

Here, we provide a brief technical overview without the use of mathematical formulations of the WT as applied to standard ISCEV ERGs [1]; readers interested in mathematical formulation can refer to the Appendix. Specifically, we describe the continuous wavelet transform (CWT) and discrete wavelet transform (DWT) and provide example application in ERGs recorded from human subjects. A short tutorial that describes the use of freely available MATLAB (Natick, MA) software that is capable of computing CWT and DWT is provided. By providing a conceptual explanation and highlighting the capacity of the CWT and DWT to reveal specific time-frequency detail, we offer a practical, readily implementable framework for leveraging wavelet analysis in ERG research and clinical interpretation.

### A High-Powered Mathematical Microscope: Continuous Wavelet Transform

Conceptually, the CWT is the convolution of a mathematical function, termed the mother wavelet, and the signal of interest. As shown in Fig. 1, the mother wavelet is shifted in time and scaled in frequency (e.g., compressed or dilated), which are often referred to as daughter wavelets. For example, the top two rows of Fig. 1 illustrate the daughter wavelet shifted in time (at time 0 and time 1). Once the daughter wavelet has completed sequential shifting and convolutions across the signal, it is scaled to new frequency (thus, new daughter wavelet), and beginning at time 0 as demonstrated in the third row. This process is iteratively repeated to assess how similar the daughter wavelets of different time and dilation are to all portions of the signal. The result is a plot of energy in the signal at a given time-frequency, as shown in the bottom row of Fig. 1 (left), and is typically referred to as a scalogram.

**Figure 1.**
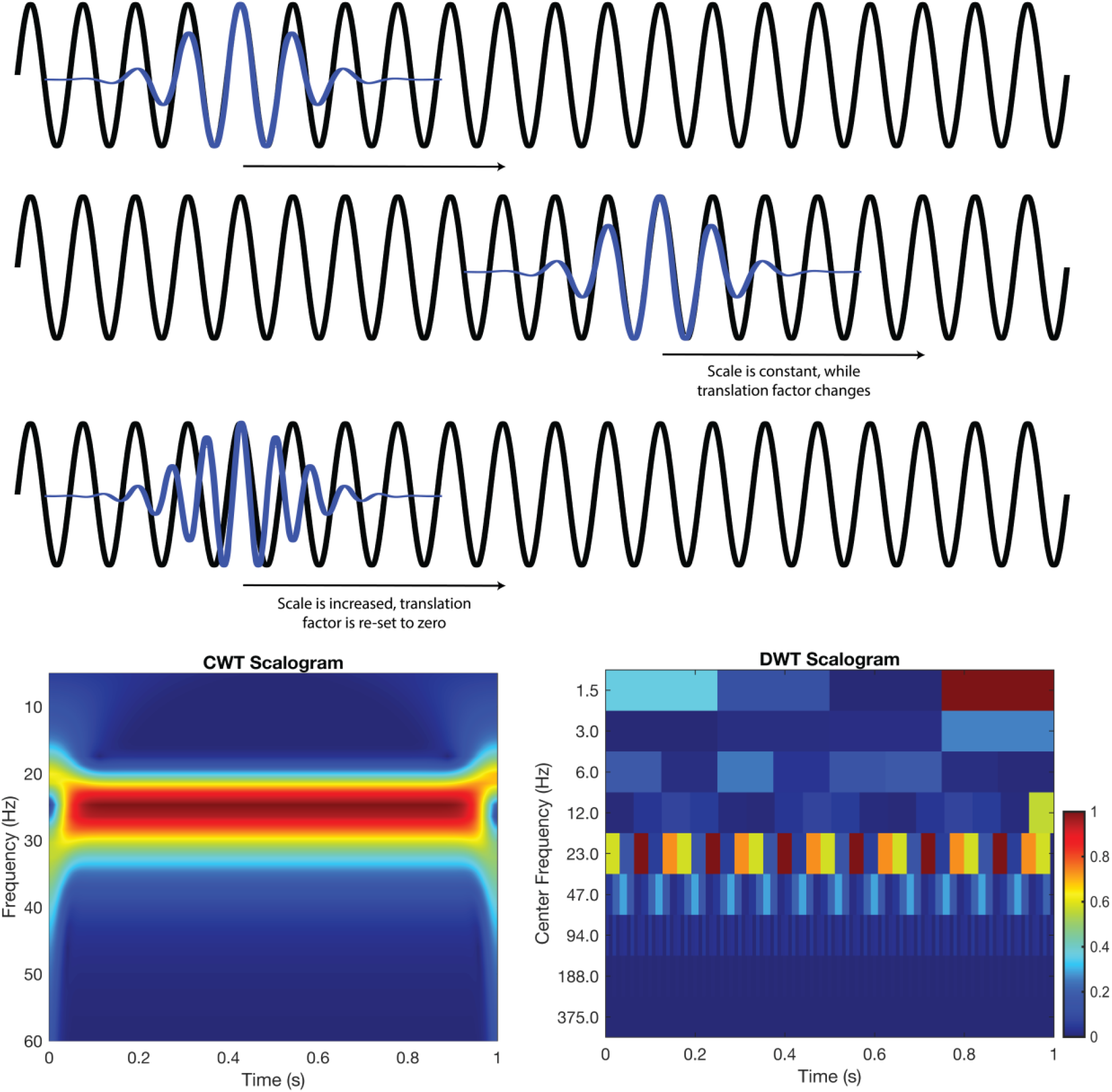
A 25 Hz sinusoid (black) is analyzed using Morlet daughter wavelets (blue). In the first two rows the daughter wavelet’s scale is fixed, giving a frequency of 25 Hz; as the daughter wavelet translates across the signal it remains perfectly in-phase, yielding high correlations and thus energy responses. In the third row, the daughter wavelets scale is dilated (scale ↑) shifting its frequency to 27 Hz, resulting in the visible mismatch and thus lower correlation/energy response. The CWT scalogram (bottom left) therefore shows a bright, continuous red band at 25 Hz with a lower energy orange band just below at 27 Hz. Similarly, the DWT scalogram (bottom right) captures the same general pattern, albeit with discrete time-frequency blocks.

### An effective balance of efficiency and detail: Discrete Wavelet Transform

Although the CWT can effectively describe the ERG, this method is computationally inefficient, as it performs redundant scales and shifts. In contrast, the DWT operates on discrete scales and dyadically down samples (powers of two), effectively balancing efficiency while capturing essential signal features. Thus, the DWT evaluates only discrete frequencies and times, rather than examining the continuous spectrum as performed by the CWT.

The DWT follows a hierarchical process, where the signal is first passed through a high pass filter, and low pass filter. The outputs of these filters are then dyadically down sampled. This ultimately yields two bands of frequency content, with the high pass filter retaining an upper band called detail coefficients, and the low pass filter retaining a lower band called approximation coefficients. The approximation coefficients again go through this process, filtering through a high pass filter and low pass filter and are then down sampled. This iterative process of filtering and down sampling continues until down sampling can no longer take place, or a predetermined level is achieved. An example of DWT decomposition can be found in Fig. 1 (bottom right). The plot is similar to the CWT (bottom left), but the energy is plotted in discrete frequency-time bins.

Not only does the DWT offer computational efficiency thanks to its hierarchical decomposition, it is an excellent compromise of all time, frequency, and time-frequency methods. This is because as each level yields a distinct set of approximation and detail coefficients, one can easily threshold coefficients from specific levels. Specifically, this can be applied to lower levels where there is unwanted noise (high frequency detail coefficients), or correct baseline drift (low frequency approximation coefficients). This multifaceted approach enables maximal physiological signal preservation while also easing the process of artifact removal. Additionally, interpreting the DWT numerical results are less subjective and straight forward compared to the CWT scalogram, as each energy index or coefficient relates to a specific time-frequency band. This discrete, block-like representation often makes it easier to compare quantitative values across multiple subjects or conditions, facilitating robust statistical analysis and clear-cut clinical or research interpretations.

For both the CWT and DWT, there are two important considerations: 1) wavelet analysis is affected by reduced reliability at the beginning and end of the signal. The wavelet cannot fully overlap the beginning and end of the signal, resulting in a region where energy estimates may be less accurate. Often, the signal is “padded” to minimize artifacts in the CWT that arise at the beginning and end of the signal. This may not be necessary for recordings that contain extended pre- or post-stimulus segments in the recording (e.g., baseline pre-stimulus recording). 2) The choice of mother wavelet is dependent on a variety of factors for both methods. In the DWT: Filter length influences temporal and spectral resolution, with shorter lengths yielding higher temporal and lower spectral resolution, and vice versa for longer length filters. Higher number of vanishing moments are better suited for removing low-order polynomial trends and capturing abrupt changes yet result in longer filter lengths. Orthogonal wavelets (Haar, Daubechies-dbN, Symlets-symN, Coiflets-coifN) yield non-redundant energy-preserving coefficients, and are thus well suited for feature extraction, while biorthogonal wavelets maintain linear-phase symmetry. Though the latter prevent phase distortion, the transform is no longer energy-orthonormal - short, fairly symmetric, orthogonal wavelets like sym2 balance these factors well for ERG analysis [19]. Like filter length modulation in the DWT, the CWT time-bandwidth product (i.e, the number of oscillatory cycles contained in the wavelet) can be adapted to improve either frequency or temporal resolution, at the cost of the other. For both DWT and CWT, selecting a mother wavelet whose form mirrors the signal’s own morphology enables the transform to better capture the underlying physiological energy patterns with maximal fidelity. Previous studies have utilized the Mexican Hat wavelet for its close match to typical ERG waveform profiles, whereas Morlet wavelets are often used to isolate high-frequency OPs in pathological ERGs [15].

### Case Study: Applying Fourier and Wavelet Methods to Healthy and CSNB ERGs

In the following section, we demonstrate the implementation and utility of WT methods in assessing ERG recordings. Code and instructions to perform this same CWT and DWT analysis are available in the Code Availability section. Analysis was performed on two right-eye example recordings from a healthy male (between 30-34 years of age), and a male (between 15-19 years of age) with complete congenital stationary night blindness (CSNB). Recordings were from the patient’s right eye using a Burian-Allen bipolar contact lens electrode. All signals were passed through a 0.3-300 Hz Butterworth band pass filter. Flash responses were sampled at 2,500 Hz, whereas the flicker recordings were sampled at 1,000 Hz. These signals are available in the Data Availability section. Rather than plotting the power spectrum, the amplitude (square root of the power) of each waveform was obtained (second column). All signals were detrended using the MATLAB detrend function, to remove DC offset prior to analysis. The study protocol was reviewed and approved by the Institutional Review Board of the University of Illinois Chicago (UIC IRB). Written informed consent was obtained from all participants, and all procedures adhered to the tenets of the Declaration of Helsinki.

Looking at the LA flicker response first, the normal tracing (Fig. 2A) shows five well-defined, evenly spaced cycles peaking near 100 µV, whereas the CSNB waveform (Fig. 2B) is flatter and reduced to ≈60 µV. The amplitude plot demonstrates a dominant 30 Hz fundamental with harmonics extending to 90 Hz in the normal eye, but in CSNB these higher-order components are significantly reduced. In the CWT the normal recording exhibits a high energy band across the entire signal, while the CSNB CWT is reduced. Likewise, the DWT reveals high energy in the 23 Hz Center Frequency, while the CSNB exhibits lower energy profiles here.

**Fig. 2.**
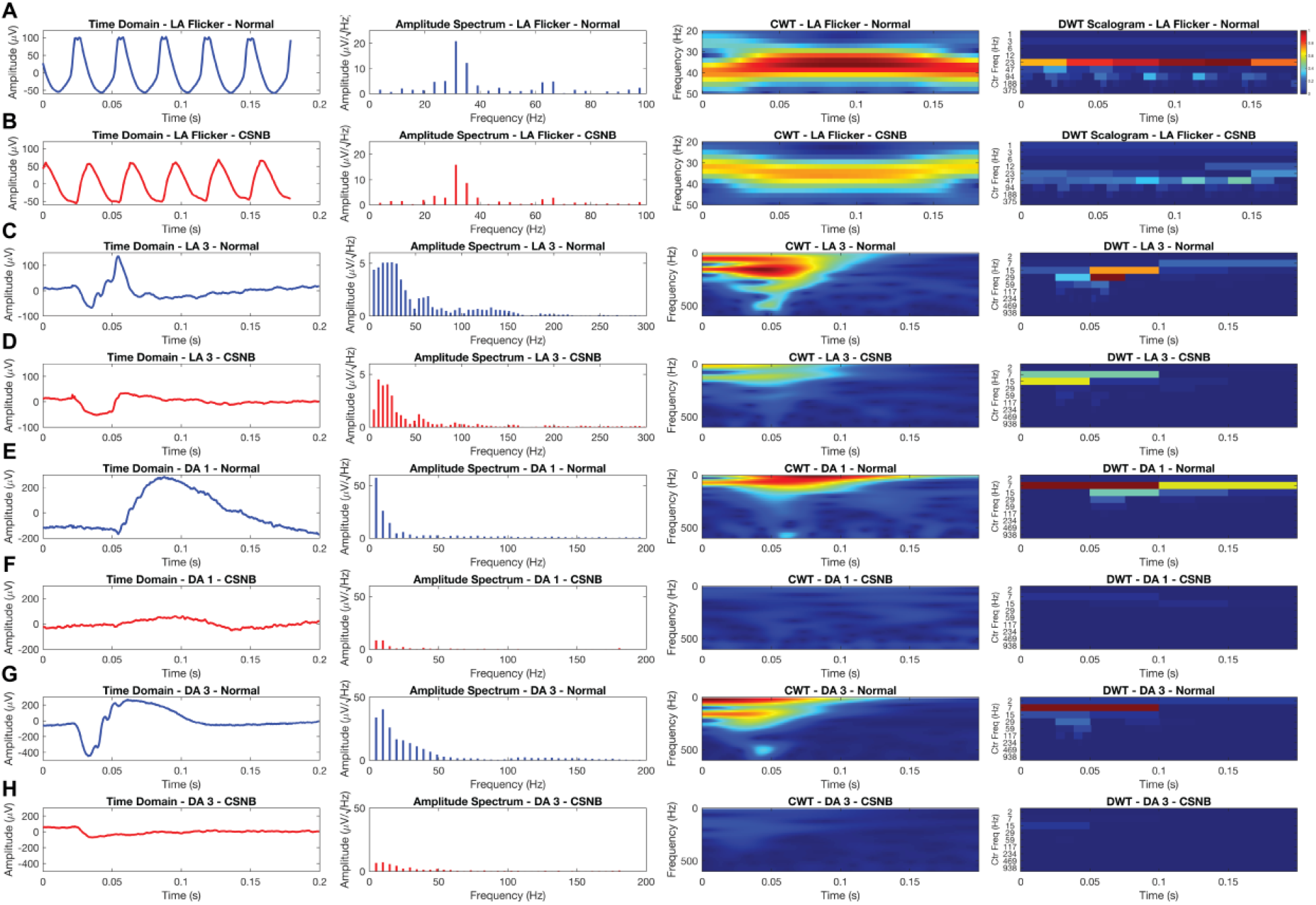
Full-field ERG recordings from a healthy subject versus a CSNB subject for the four ISCEV standard recordings: (A, B) LA Flicker, (C, D) LA 3.0 cd·s/m^2^, (E, F) DA 0.01 cd·s/m^2^, and (G, H) DA 3.0 cd·s/m^2^. Each row presents the time-domain waveform, corresponding amplitude spectrum, and both continuous wavelet transform (CWT) and discrete wavelet transform (DWT) scalograms. Differences in amplitude, peak frequency content, and localized time-frequency energy highlight the functional impact of CSNB across cone- and rod-dominated responses.

In the LA 3 flash, the normal tracing (Fig. 2C) shows a sharp negative deflection followed by a positive wave, while the CSNB recording (Fig. 2D) is attenuated and delayed. Amplitude spectrums demonstrate higher valued responses in the normal recording but reduced and shifted amplitude response in CSNB. Following this, the CWT displays high energy < 100 Hz between 0.04-0.08 s in the normal recording while the CSNB recording exhibits most of its response, albeit attenuated, < 50 Hz. However, when evaluating the DWT, the normal subject exhibits most of its energy in the 29 Hz center frequency band from 0.05-0.075 s, while the CSNB is attenuated here, but presents with a greater energy in the 7 and 15 Hz center frequency band in the first half of the signal. For the DA 1 traces, the normal response (Fig. 2E) presents a clear biphasic pattern with an early trough and positive peak, while the CSNB trace (Fig. 2F) exhibits much lower amplitude. The amplitude spectrum of the normal contains a wider 10-40 Hz envelope, while the CSNB amplitude is largely diminished, and only perceptible < 15 Hz. In the CWT, the normal recording shows discrete power bursts aligned with its main peaks, whereas the CSNB distribution is entirely absent in comparison. Similarly, the DWT exhibits high energies in the 7 Hz center frequency band for the entire signal while the CSNB response is relatively absent.

Lastly in the DA 3 responses, the normal waveform (Fig. 2G) features a pronounced negative trough followed by a large positive peak around 0.07 s, while the CSNB trace (Fig. 2H) is almost entirely flat. Amplitude spectrum reveals distinct 5-50 Hz peaks in the normal but negligible energy in CSNB. The CWT captures two distinct high energy areas in the first half of the signal that are almost completely absent in the CSNB. Likewise, the DWT demonstrates concentrated energy in the first half of the 7 Hz center frequency band in the normal that is largely absent in CSNB, demonstrating the marked loss of dark-adapted responsiveness typically seen in CSNB.

## Conclusion

As demonstrated, advanced signal processing techniques like the CWT and DWT provide qualitative and quantitative insights for ERG analysis. While time-domain features offer some pathological insight, a complete picture can be obtained with these additional time-frequency based measures. However, no single method is superior to another, and the analytical choice of approach is dependent on a variety of factors including stimulus, pathology, or feature of interest. Ultimately, integrating wavelet-based techniques into routine ERG analysis may significantly enhance diagnostic accuracy and provide deeper insight into retinal function and pathology.

## Data Availability

The data and code utilized in this study is made available in the following GitHub repository: https://github.com/yshwetar/ERG-Signal-Processing/tree/main.

## Acknowledgements

YJS and MAH would like to thank the University of North Carolina at Chapel Hill, School of Medicine Genetics Department for funding and support.

## Author Contributions

Yousif Shwetar conceived and wrote the first draft of the manuscript. David Lalush, Brett Jeffrey, Jason McAnany, and Melissa Haendel contributed critical revisions and guidance. All authors commented on previous versions of the manuscript and approved the final version.

## Declarations Data Availability

The data utilized in this study is made available in the following GitHub repository.

## Code Availability

Code to execute discussed analysis is available in the following GitHub repository.

## Ethics Approval

Approved by the Institutional Review Board of the University of Illinois Chicago (UIC IRB).

## Consent to Participate

Written informed consent was obtained from all participants.

## Consent to Publish

Participants provided consent for publication of anonymized data.

## Informed Consent

Written informed consent was obtained from all participants.

## Statement of Human Rights

All procedures conformed to the Declaration of Helsinki.

## Statement on the Welfare of Animals

Not applicable.

## APPENDIX

### Continuous Wavelet Transform

The continuous wavelet transform (CWT) is defined by:

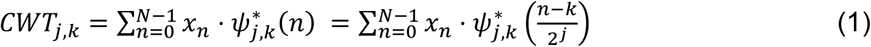

Where:

- *x*_*n*_ is the original signal,
- *ψ*(*n*) is the mother wavelet,
- *j* and *k* control scaling (frequency) and shifting (time), respectively
- * denotes complex conjugation.

Each mother wavelet *ψ* has a central frequency *F*_*c*_. Scaling by a factor *a* yields a daughter wavelet with a pseudo-frequency *F*_*a*_:

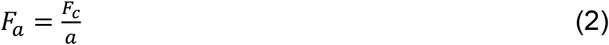

In practice, *a* = 2^*j*^. The local wavelet energy is:

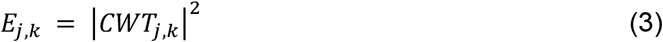

The mother wavelet’s central frequency *F*_*c*_ defines its fundamental sinusoidal structure, so when you scale and slide (*j* and *k*, respectively) *ψ*(*n*) across the signal *x*_*n*_, those segments of *x*_*n*_ that most closely match *ψ*(*n*) at that pseudo frequency exhibit high correlation (and hence high energy, *E*_*j,k*_), whereas poorly matched regions produce near-zero coefficients.

#### Discrete Wavelet Transform

The DWT is defined as follows:

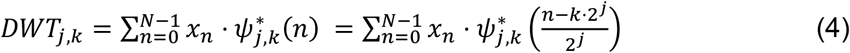

In practice, the DWT is implemented by passing the original signal through the following steps:

1. A high-pass filter and low-pass filter yielding detail and approximation coefficients.
2. Both sets of coefficients are then down sampled by a factor of two.
3. Approximation coefficients are selected and iteratively processed through steps 1-3 until decomposition can no longer takes place.

Mathematically, each decomposition level is defined as *j*:

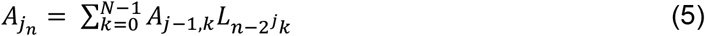

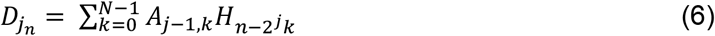

Energy of these approximation and detail coefficients are calculated as:

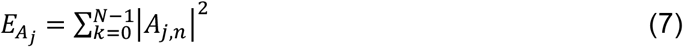

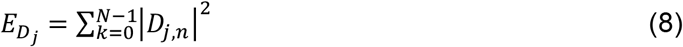

